# A systematic review and meta-analysis of inpatient mortality associated with nosocomial and community COVID-19 exposes the vulnerability of immunosuppressed adults

**DOI:** 10.1101/2021.07.10.21260306

**Authors:** Mark J. Ponsford, Tom JC Ward, Simon Stoneham, Clare M. Dallimore, Davina Sham, Khalid Osman, Simon Barry, Stephen Jolles, Ian R. Humphreys, Daniel Farewell

## Abstract

**Background:** Little is known about the mortality of hospital-acquired (nosocomial) COVID-19 infection globally. We investigated the risk of mortality and critical care admission in hospitalised adults with nosocomial COVID-19, relative to adults requiring hospitalisation due to community-acquired infection.

**Methods:** We systematically reviewed the peer-reviewed and pre-print literature from 1/1/2020 to 9/2/2021 without language restriction for studies reporting outcomes of nosocomial and community-acquired COVID-19. We performed a random effects meta-analysis (MA) to estimate the 1) relative risk of death and 2) critical care admission, stratifying studies by patient cohort characteristics and nosocomial case definition.

**Results:** 21 studies were included in the primary MA, describing 8,246 admissions across 8 countries during the first wave, comprising 1517 probable or definite nosocomial COVID-19, and 6729 community-acquired cases. Across all studies, the risk of mortality was 1.31 times greater in patients with nosocomial infection, compared to community-acquired (95% CI: 1.01 to 1.70). Rates of critical care admission were similar between groups (Relative Risk, RR=0.74, 95% CI: 0.50 to 1.08). Immunosuppressed patients diagnosed with nosocomial COVID-19 were twice as likely to die in hospital as those admitted with community-acquired infection (RR=2.14, 95% CI: 1.76 to 2.61).

**Conclusions:** Adults who acquire SARS-CoV-2 whilst already hospitalised are at greater risk of mortality compared to patients admitted following community-acquired infection; this finding is largely driven by a substantially increased risk of death in individuals with malignancy or who had undergone transplantation. These findings inform public health and infection control policy, and argue for individualised clinical interventions to combat the threat of nosocomial COVID-19, particularly for immunosuppressed groups.

Systematic review registration: PROSPERO CRD42021249023

## 1 Introduction

Health-care-associated infections represent an enduring and serious threat to patient safety (1,2), and are estimated to cost the National Health Service (NHS) £1 billion each year (3). The transmission of respiratory viruses such as influenza in the healthcare environment are a well-recognized cause of significant morbidity and mortality at the individual patient level (4), however less is known regarding the significance of in-hospital (nosocomial) transmission of the novel pandemic coronavirus SARS-CoV-2 causing COVID-19 (5). Since its emergence in 2019, COVID-19 has placed enormous pressure on health-care systems worldwide. Limited availability of testing, asymptomatic infections, and an evolving understanding of routes of transmission have led to the exposure of potentially vulnerable uninfected patients in the health-care setting (6).

The first and only rapid literature review and meta-analysis conducted to date on nosocomial COVID-19 in hospitalised individuals was published in April 2020, early in the course of the pandemic, and included only 3 studies reporting prevalence (7). The UK COVID-19 Clinical Information Network (CO-CIN) estimated 31,070 nosocomial COVID-19 infections occurred in England between February and July 2020, but made no assessment of the risk of mortality (8). We recently reported our initial experience from the first wave of the COVID-19 pandemic across the nation of Wales, using data collected from 2508 hospitalised adults (9). In this observational study, inpatient mortality rates for nosocomial COVID-19 ranged from 38% to 42% and were consistently higher than participants with community-acquired infection (31% to 35%) across a range of possible case definitions. Whilst supported by other studies (10,11), this finding contrasts with several earlier reports suggesting that nosocomial COVID-19 infection is associated with a similar risk of inpatient mortality to community acquired infection (12–14).

It is well known that individuals with pre-existing health conditions particularly ischemic heart disease, diabetes, hypertension and immunosuppression (15–17), as well as older and frailer individuals (18), are at increased risk of death from SARS-CoV-2. Such individuals are also likely to be over-represented in inpatient cohorts (19). Together, this suggests a robust assessment of the burden of mortality is urgently needed to examine the risk to patients, identify vulnerable cohorts, and direct policies to ensure improvement. We therefore performed a systematic review and meta-analysis of published and pre-print studies reporting mortality associated with probable and definite nosocomial SARS-CoV-2 outbreaks during the first wave of the COVID-19 pandemic. Our primary aim was to compare case fatality rates of nosocomial and community-acquired COVID-19 cases within hospitalised adults. Our secondary aims were to assess the variation in risk of mortality between patient sub-groups, the relative risk of critical care admissions, and to probe the risk of bias associated with these reports. Together, this provides a timely insight to the global burden of hospital-acquired COVID-19, and highlights key patient groups at elevated risk of mortality due to nosocomial exposure. These findings inform public health policy, and argue for enhanced infection control alongside and access to clinical interventions to combat the threat of nosocomial COVID-19.

## 2 Methods

We followed the Preferred Reporting Items for Systematic Reviews and Meta-Analyses (PRISMA) 2020 (20). The study protocol was prospectively registered with Prospero (CRD42021249023), having first confirmed no similar reviews were underway.

### 2.1 Eligibility criteria

#### 2.1.1 Participants

Studies of hospitalised adults (≥16 years) within acute or long-term healthcare settings, excluding care or residential homes. We specifically focused on outcomes for hospitalised adults and excluded outcomes from health care workers with nosocomial infection, as the latter has been recently evaluated (21).

#### 2.1.2 Exposures

We included any implicit or explicit case definition of probable or definite nosocomial acquisition as defined by the study authors, considering these further in sensitivity analyses. Patients where COVID-19 origin was unclassified were excluded. Implementation of universal screening of patients and healthcare workers, and changes to personal protective equipment have recently been reported in detail elsewhere (22) and were not further considered.

#### 2.1.3 Comparators

The number and outcome of adults hospitalised with community-acquired SARS-CoV-2 within the same study setting.

#### 2.1.4 Outcomes

The primary outcome was mortality of nosocomial SARS-CoV-2 infections in hospitalised adult patients and community-acquired SARS-CoV-2 infection. Secondary outcomes included rates of critical care admission, and qualitative analysis of case definitions, study timing, and variation in reporting by country of origin.

#### 2.1.5 Study design

Observational case series and cohort studies were included, provided they reported an outbreak of nosocomial SARS-CoV-2 (defined as ≥2 patients with likely nosocomial infection) within the hospital setting. Case reports with a single participant (high risk of bias, unable to assess proportion/risk), exclusively outpatient populations (e.g. dental practice), and non-patient populations (e.g. healthcare workers only) were therefore excluded.

### 2.2 Search strategy to identify studies

#### 2.2.1 Database search strings

Ovid Medline, Embase, and the Social Policy & Practice databases and MedRvix.org were searched from 1/1/2020 to 9/2/2021. A search string was designed that included the following concepts: [SARS-CoV-2 OR sars-cov 2 OR COVID-19 OR covid 19 OR 2019-nCoV or “COVID-19”] AND [nosocomial OR hospital-acquire* or nosocomial-acquire* OR cross infection].

#### 2.2.2 Restriction on publication type

No restrictions by language were imposed, and Google Translate was used to review full text documents where required. In addition to considering full-text articles, publications available as abstract only were included if they contained sufficient information to inform the primary outcome.

#### 2.2.3 Study selection and screening

Five clinicians (MJP, TJCW, SS, DS, KO, CD) independently screened titles and abstracts against inclusion criteria using Rayyan (23). MJP retrieved the full-texts, and with TJCW and SS screened these for inclusion. Conflicts were resolved by consensus. The selection process is outlined in the PRISMA flow diagram (Figure 1).

**Figure 1:**
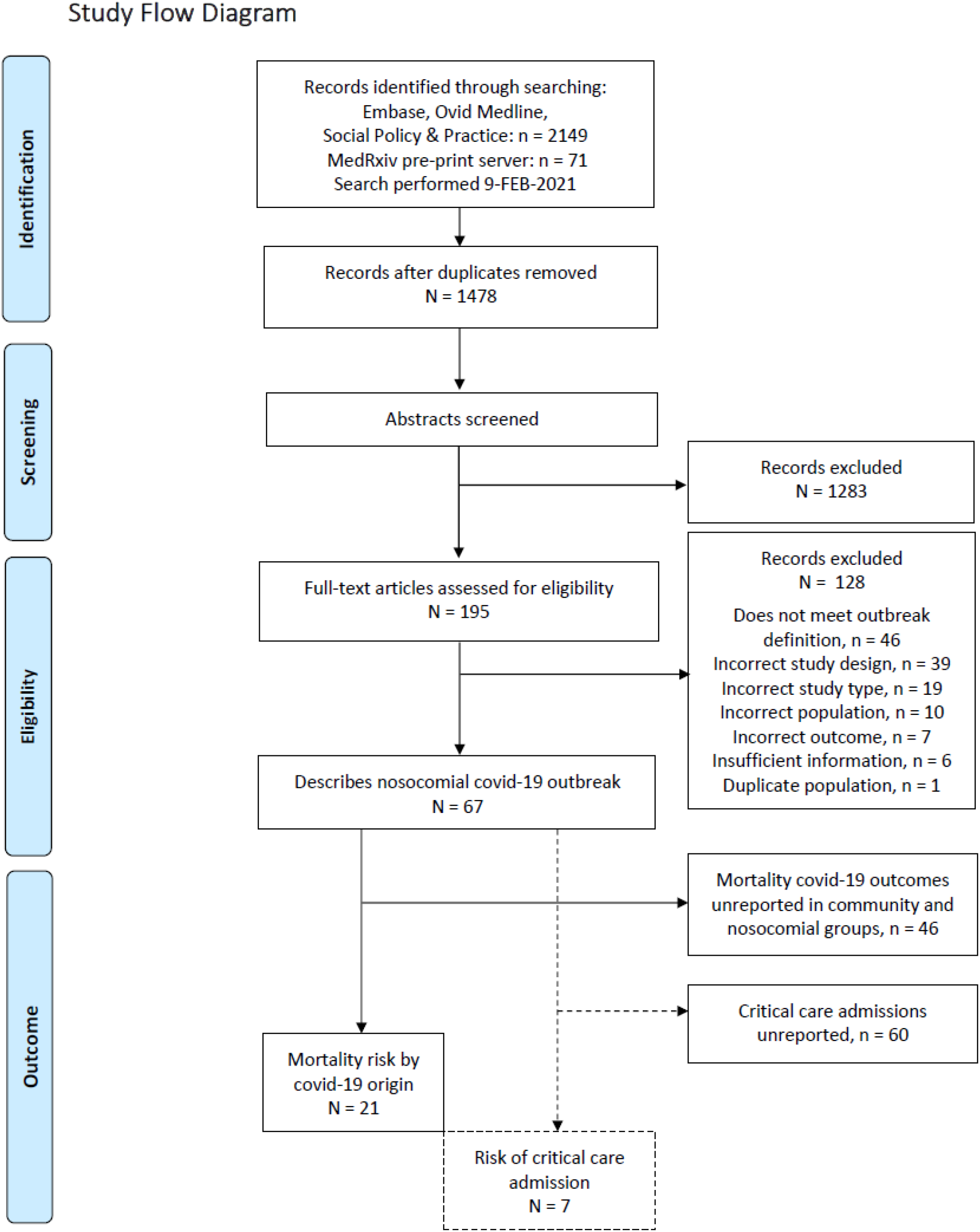
PRISMA Study Flow Diagram.

### 2.3 Data extraction

Data was extracted using a pre-defined spreadsheet with fields as presented in Table 1, and cross-checked for accuracy and completeness by a second reviewer. COVID-19 case diagnosis rates by country were retrieved from https://ourworldindata.org/coronavirus-source-data on 6th April 2021.

**Table 1:**
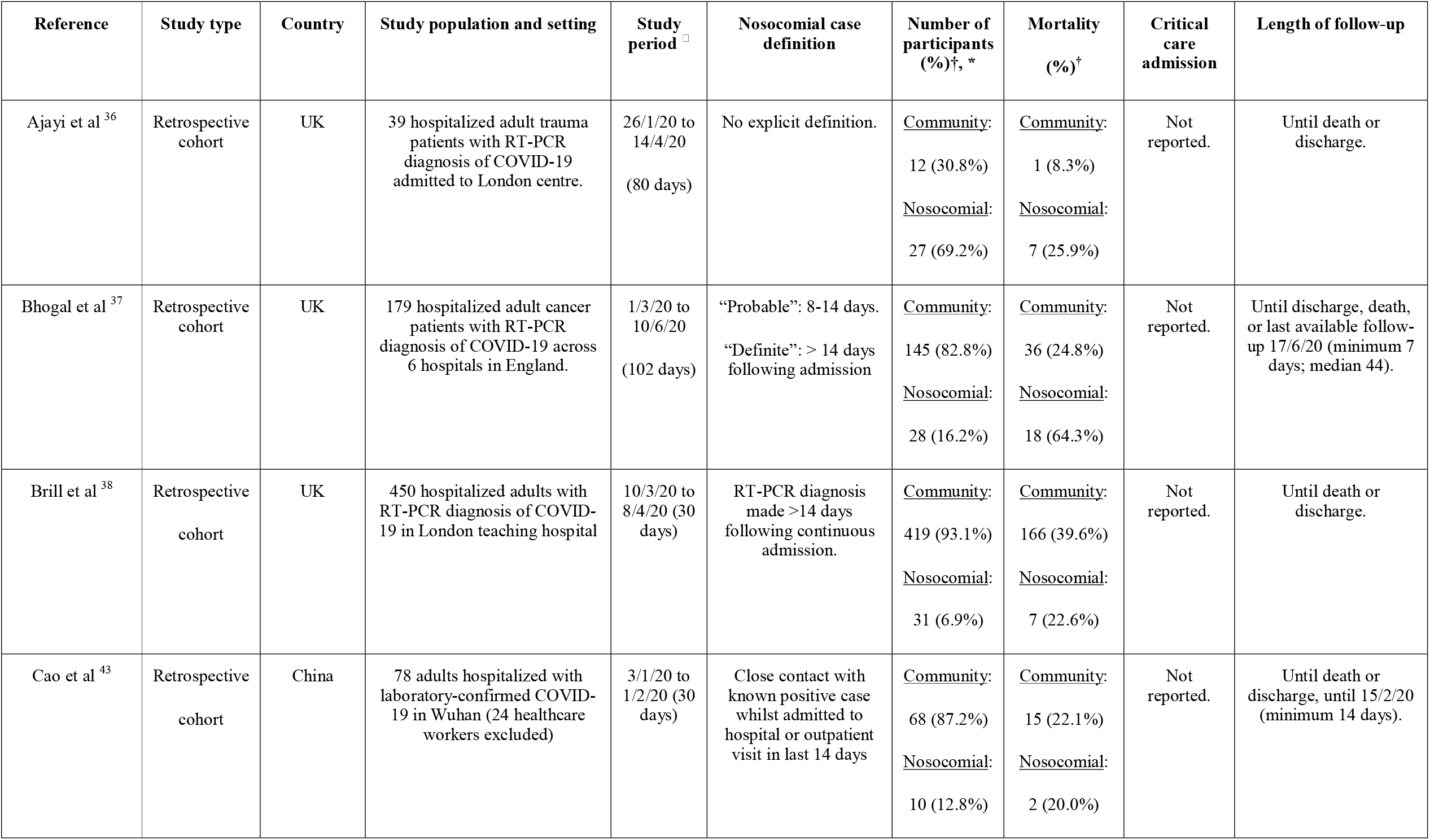
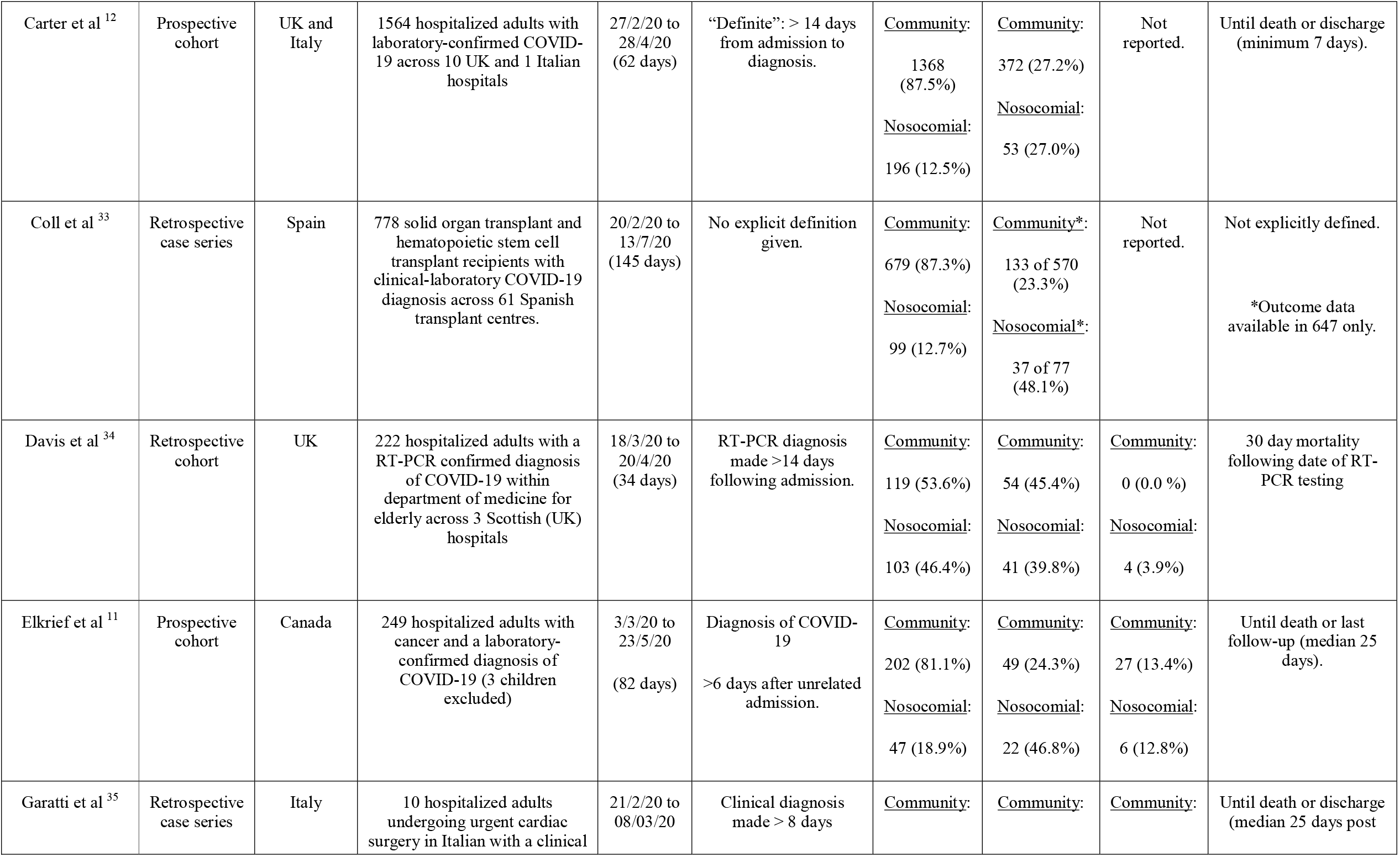
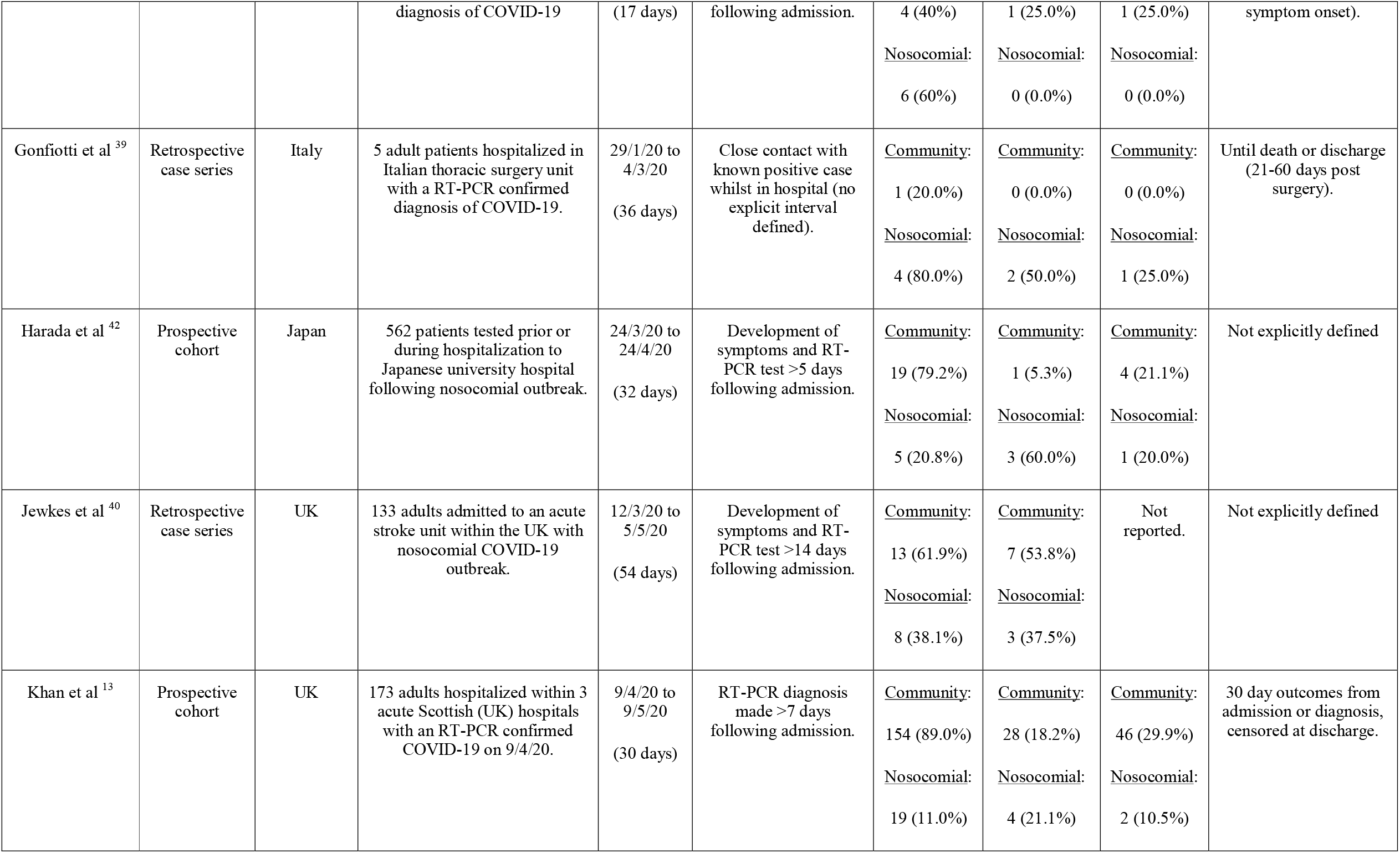
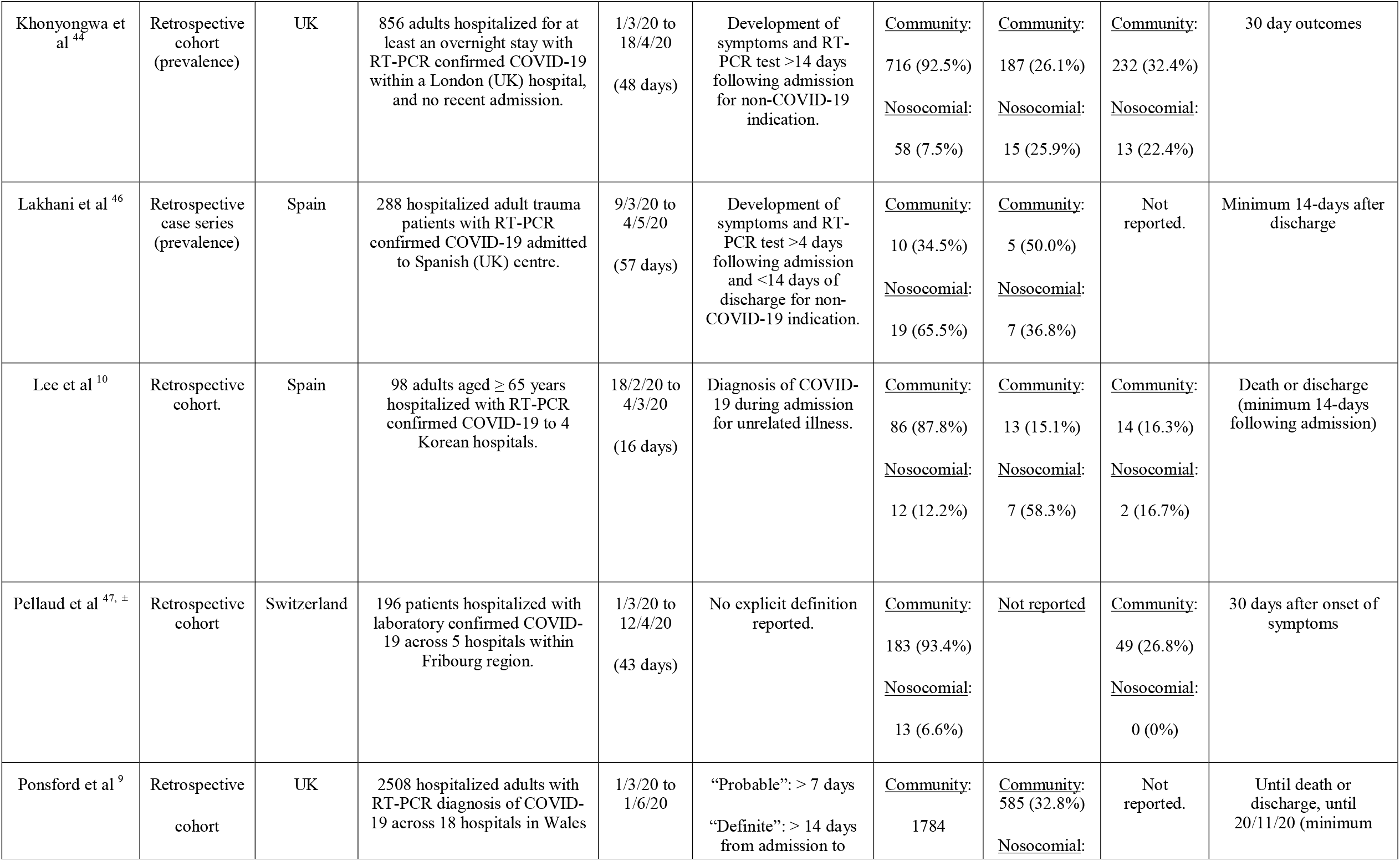
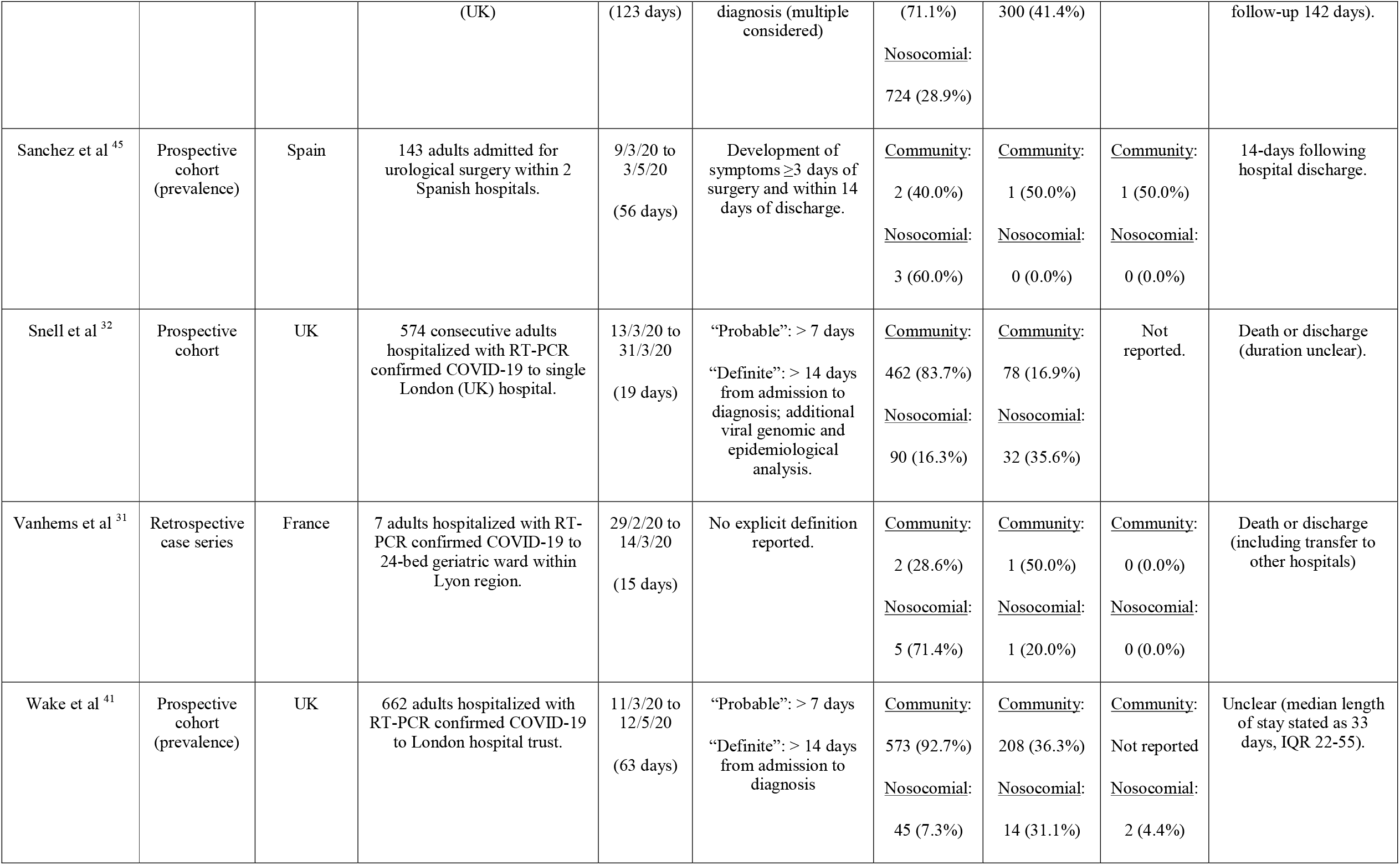
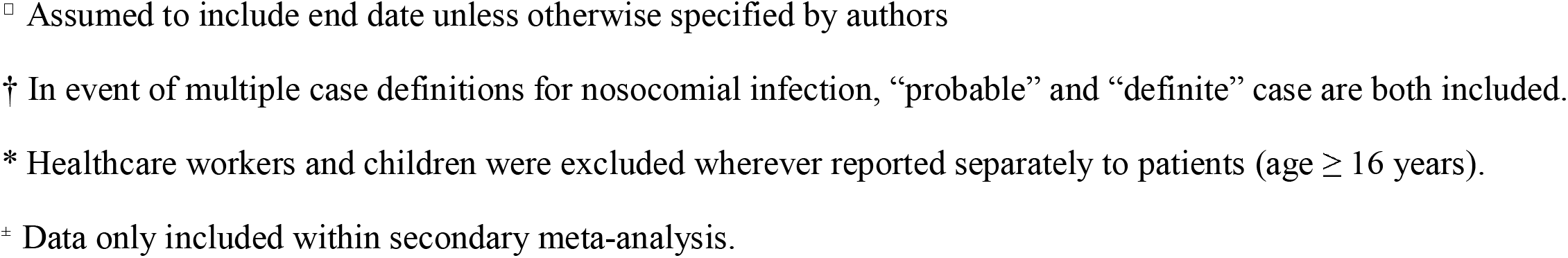
Evidence Summary Table.

### 2.4 Assessment of risk of bias

Formal risk of bias on a study and outcome level were conducted using the Newcastle Ottawa Score (NOS) for cohort studies and Joanna Briggs Institute (JBI) tools for case series and prevalence studies (24), as recommended by the National Institute for Clinical Excellence (NICE) (25). Assessment was performed by 2 independent reviewers, with arbitration with a third as required. We defined adequate follow-up as ≥28 days, or complete follow-up until death or discharge, to account for the potential unequal time points in disease course at study entry between community and nosocomial patients. We considered principal areas likely to introduce bias, indicated by * in Tables 2-4, equating to a minimum score of 5 across tools. Briefly, these assessed quality of selection: a) representativeness of the average nosocomial covid-19 case within the patient group, based on study design, b) ascertainment bias – requiring evidence of methods to mitigate this, c) sufficient description of study subjects and case definition – requiring an explicit nosocomial case definition given and applied; and quality of outcome assessment: a) sufficient follow-up, and b) adequacy of follow-up – requiring sufficient participants to have reached the pre-specified outcome at time of reporting.

**Table 2:** Risk of Bias Assessment - cohort studies (n=8)

| Study<br>Author | Domain<br>1* | Domain<br>2* | Domain<br>3* | Domain<br>4 | Domain<br>5 | Domain<br>6 | Domain<br>7* | Domain<br>8* | Total<br>Score |
| --- | --- | --- | --- | --- | --- | --- | --- | --- | --- |
| Ajayi et al | 1 | 1 | 1 | 0 | 0 | 1 | 0 | 0 | 4 |
| Brill et al | 1 | 1 | 1 | 0 | 0 | 1 | 1 | 0 | 5 |
| Lee et al | 1 | 1 | 1 | 0 | 0 | 1 | 0 | 1 | 5 |
| Bhogal et al | 1 | 1 | 1 | 0 | 1 | 1 | 1 | 0 | 6 |
| Elkrief et al | 1 | 1 | 1 | 0 | 1 | 1 | 1 | 0 | 6 |
| Carter et al | 1 | 1 | 1 | 0 | 1 | 1 | 1 | 1 | 7 |
| Khan et al | 1 | 1 | 1 | 0 | 1 | 1 | 1 | 1 | 7 |
| Ponsford et al | 1 | 1 | 1 | 0 | 1 | 1 | 1 | 1 | 7 |

**Table 3:**
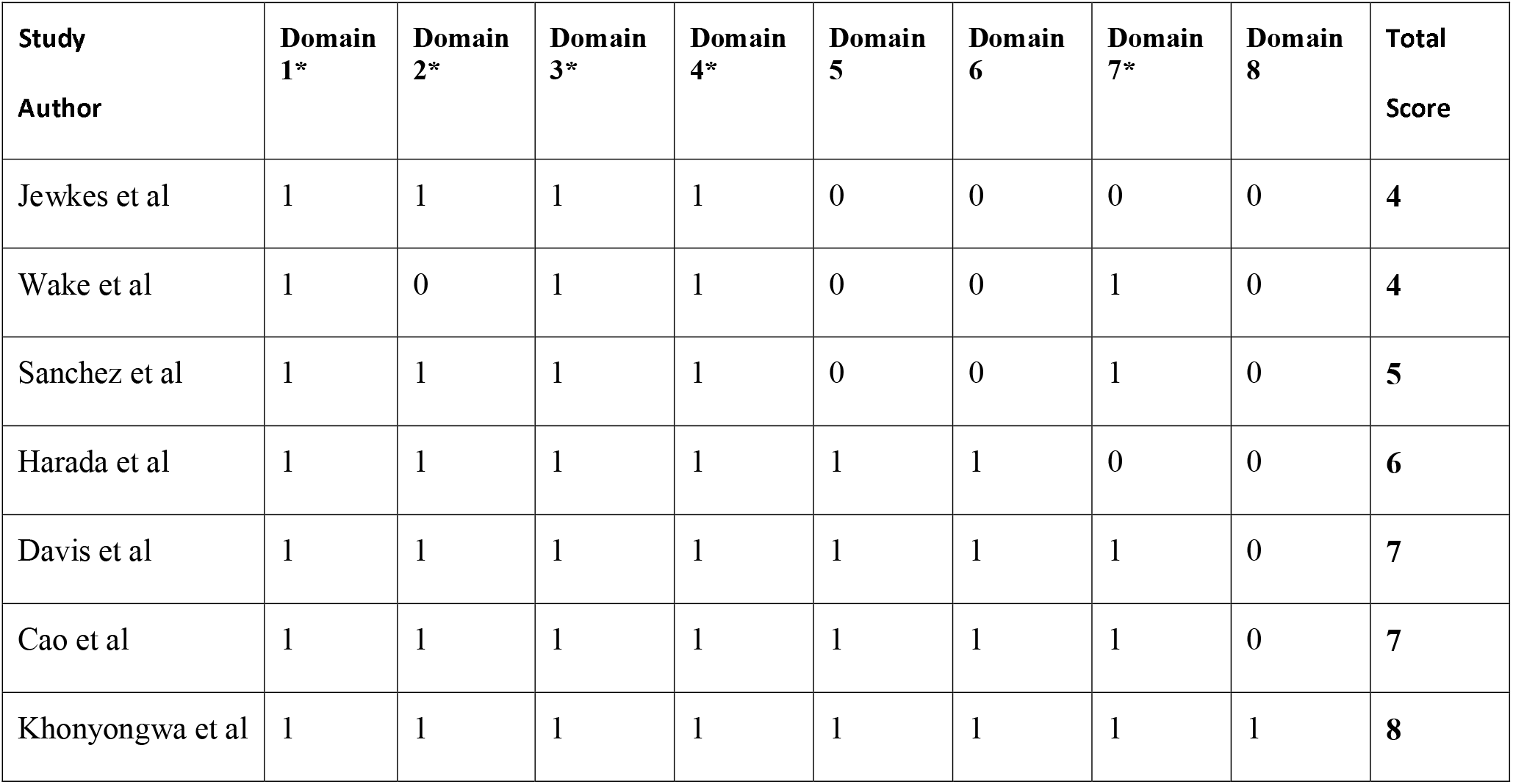
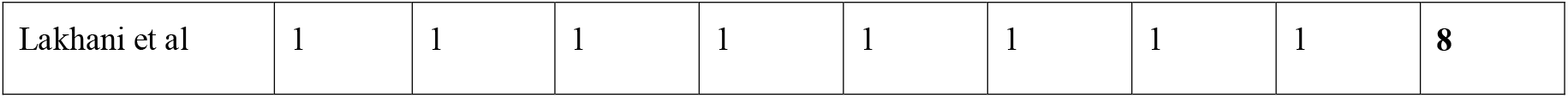
Risk of bias assessment - prevalence studies (n=8)

**Table 4:** Risk of bias assessment - case series (n=5)

| Study | Domain 1* | Domain 2* | Domain 3 | Domain 4* | Domain 5 | Domain 6 | Domain 7 | Domain 8* | Domain 9* | Domain 10 | Total Score |
| --- | --- | --- | --- | --- | --- | --- | --- | --- | --- | --- | --- |
| Vanhems et al | 1 | 0 | 0 | 0 | 0 | 1 | 1 | 1 | 0 | 0 | 4 |
| Snell et al | 1 | 1 | 1 | 1 | 0 | 0 | 0 | 1 | 1 | 0 | 6 |
| Coll et al | 1 | 0 | 1 | 0 | 0 | 1 | 1 | 0 | 1 | 1 | 6 |
| Gonfiotti et al | 0 | 1 | 1 | 1 | 0 | 1 | 1 | 1 | 1 | 0 | 7 |
| Garatti et al | 1 | 0 | 0 | 1 | 1 | 1 | 1 | 1 | 1 | 1 | 8 |

### 2.5 Data Analysis

Analysis was performed using R version 4.0.2 in RStudio (Version 1.3.959, R Foundation, Vienna, Austria) using the metafor package. Full details can be found within the online supplementary methods. Briefly, a random effects model was used to compare relative risk of mortality and ICU admission between patients with community-acquired and nosocomial COVID-19. Full details of the statistical methods used are available at https://cran.r-project.org/web/packages/metafor/metafor.pdf. Residual maximum likelihood (REML) was used to estimate the heterogeneity variance (τ^2^) (26). We conducted subgroup analyses based on classifications agreed by the reviewers reflecting the cohort best represented by the studies, i.e. in cohorts that were clinically and methodologically similar (27). Cochrane’s Q-test and I^2^ were used to assess the degree of inconsistency across studies (28,29). Two-sided statistical significance was set at p<0.05. We conducted the following pre-specified sensitivity analyses:

- 1: Studies providing an explicit definition of nosocomial acquisition
- 2: Studies providing outcomes associated with a standardised >14 day definition for ‘definite’ nosocomial covid-19
- 3A: Excluding studies with a higher risk of bias (indicated by total quality score <5)
- 3B: Fulfilling all 5 core study quality domains (indicated by * within tables 2-4).
- 4: Excluding studies with imputed data (i.e. 0.5 used in place of zero-count cells)

Additional data visualization was performed in R using the ggplot2 package.

### 2.6 Reporting bias assessment

Funnel plot and Egger’s test were used to assess for potential publication bias, supported by qualitative evaluation.

### 2.7 Certainty assessment

The certainty of evidence was rated using the Grading of Recommendations, Assessment, Development and Evaluations (GRADE) approach (30) using the GRADEPro online tool, https://gradepro.org/.

## 3 Results

### 3.1 Study selection and characteristics

We screened a total of 1478 unique abstracts and reviewed 195 full texts to identify 67 studies describing hospital nosocomial COVID-19 outbreaks. Principal reasons for study exclusion are shown in Figure 1. A further 48 studies were excluded as they did not report mortality within both community and nosocomial-acquired COVID-19 patient groups. This left 21 studies for primary meta-analysis (9–13,31–46), summarised in Table 1, with both retrospective (n=14) and prospective (n=7) study designs including a range of medical and surgical patient populations. Together, these described 8246 hospitalised adults admitted between 1^st^ March 2020 and 13^th^ July 2020 across 7 countries, comprising 1517 (18%) probable or definite nosocomial COVID-19 and 6729 (82%) community-acquired cases. Overall mortality was 30.5% (2516/8246), with 575 deaths attributed to nosocomial COVID-19 (37.9% mortality rate) and 1941 (28.9% mortality rate) to community-acquired COVID-19. An additional study reporting the critical care admissions but without mortality by probable-nosocomial origin was identified, and is included Table 1 (47).

### 3.2 Study timing in pandemic course and availability of universal RT-PCR testing

We explored the timing of patient identification within these reports relative to national COVID-19 diagnosis rates based on publicly available data within the UK (Figure 2), and wider countries (Supplementary S2). All included studies dealt with the initial wave of the pandemic. Consistent with the early timing of these reports, no studies reported the use of universal RT-PCR screening of patients in prior to or during admission from the outset of the study, outside of the setting nosocomial outbreaks.

**Figure 2:**
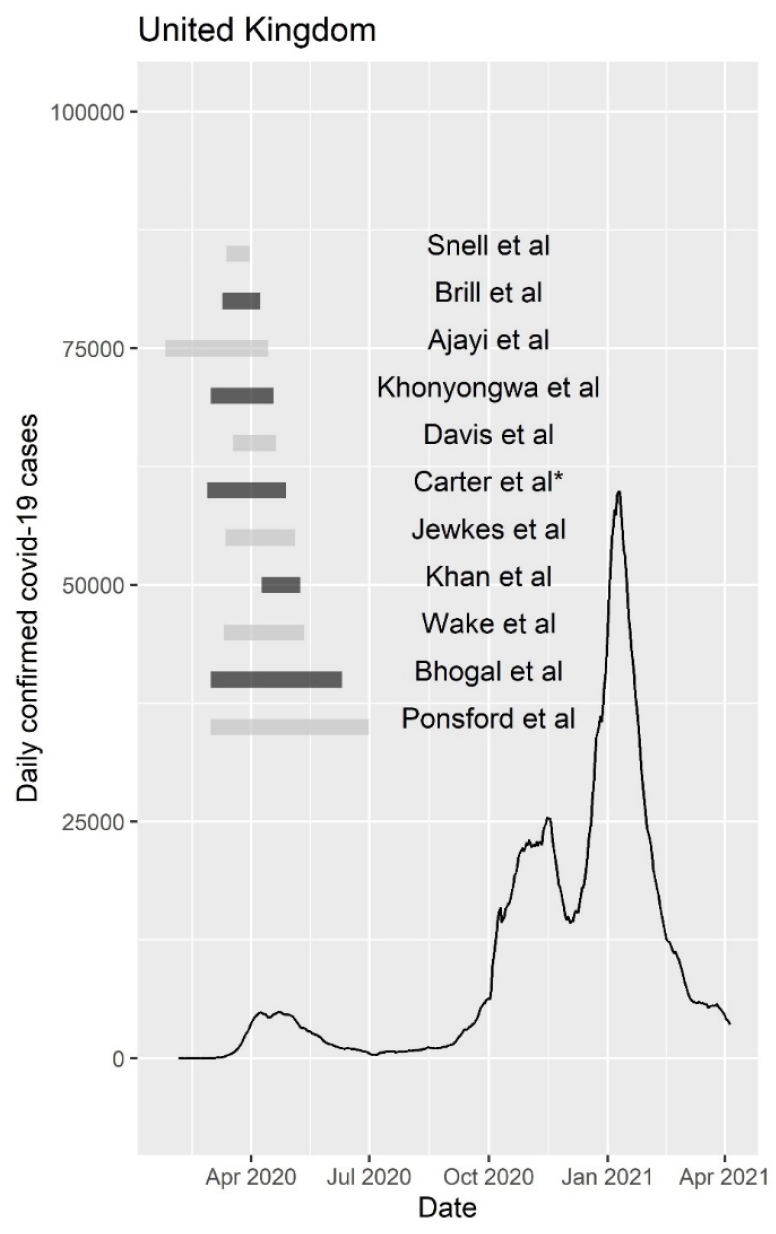
Timing of UK studies relative to national COVID-19 rates. Plot showing the timing of individual studies included within the primary meta-analysis reporting patients within the United Kingdom (UK), relative to national daily COVID-19 case diagnosis rates January 2020 and April 2021. * The study by Carter et al is included here as 10/11 hospital sites were within the UK.

### 3.3 Case definitions

A positive reverse transcription polymerase chain reaction (RT-PCR) SARS-CoV-2 result was explicitly used as primary method of diagnosis in 17/21 studies included in the mortality meta-analysis (76%), supported by clinical-radiological features (12,33,41), or based upon laboratory-based diagnosis (potentially including serology) (43,45). As shown in Table 1, a range of case definitions were employed to distinguish community-acquired and nosocomial COVID-19. A fixed interval between admission and diagnosis was employed in 14/21 (62%) ranging from >2 days (45) to >14 days (12), supplemented by additional patient-level clinical data (41) and viral whole genome sequencing (32). Seven studies primarily employed epidemiological nosocomial definitions, for instance a history of close contact with positive cases (n=3, (31,39,43)), or the absence of symptoms on admission with subsequent positive test (n=2, (10,35)). Two studies gave no explicit nosocomial case definition (33,36). Four studies (19%) explicitly considered patients who had been recently discharged.

### 3.4 Risk of bias in studies

We screened study quality through self-identified use of reporting standards. Three (14%) reports referenced the STrengthening the Reporting of OBservational studies in Epidemiology (STROBE) statement (9,12,36). Tables 2-4 show the formal risk of bias assessments. Overall, 17/21 (81.0%) achieved a total score of 5 or more. Utilising our more stringent assessment of study quality across all core domains (indicated by *) only 9/21 (43.0%) were identified, with 80% case series, 62.5% cohort, and 37.5% of prevalence rated at high risk of bias.

### 3.5 Meta-analysis of mortality in patients with nosocomial relative to community-acquired COVID-19

Meta-analysis using a random effects model is shown in Figure 3. Across 21 studies, the risk of mortality was 1.31 (95% CI: 1.01 to 1.70) times greater in patients with probable or definite nosocomial infection, compared to those admitted with community-acquired COVID-19. Substantial heterogeneity was evident between the included studies (Q= 76.3, p < 0.0001; I^2^ = 82.3%, 95% CI: 62.0 to 94.6%). We performed sub-grouping by patient cohort characteristics, including an immunosuppressed sub-group comprising 3 studies reporting outcomes from adult recipients of solid-organ or bone marrow transplants, or with a diagnosis of haematological or solid-organ cancers. These 1069 patients (152 nosocomial, 917 community-acquired) showed an elevated risk of death associated with nosocomial COVID-19, relative to those with community-acquired infection: RR= 2.14, 95% CI: 1.76 to 2.61. This effect appeared consistent across the 3 studies, but with considerable uncertainty associated with estimates of heterogeneity (Q= 1.24, p= 0.54; I^2^ = 0.00%, 95% CI: 0.00 to 96.6%). General medical and geriatric admissions were also suggesting of an increased risk of mortality with nosocomial COVID-19 (RR 1.14 and 1.35 respectively), but did not reach statistical significance.

**Figure 3:**
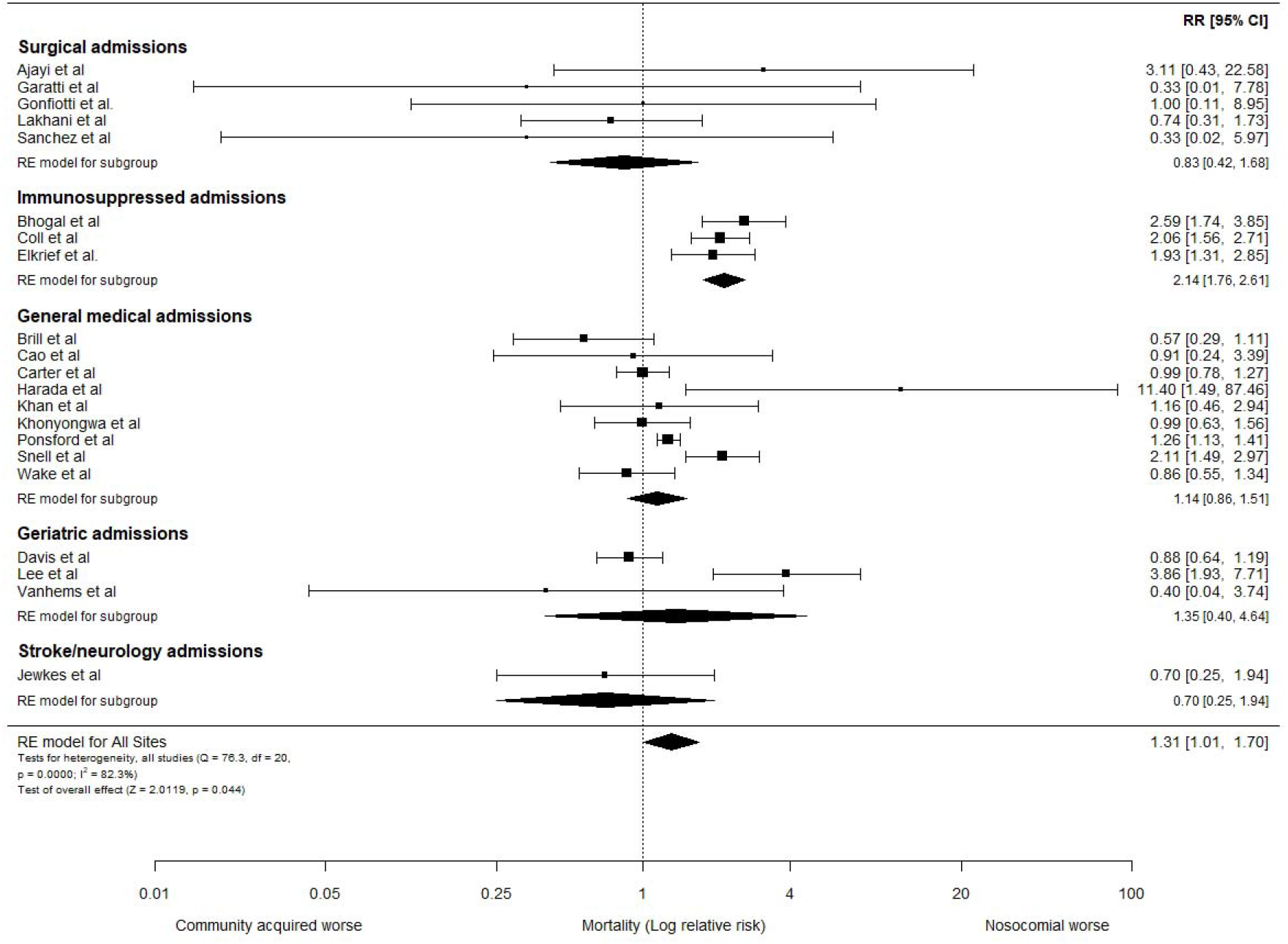
Relative risk of mortality in hospitalized adults with nosocomial and community-acquired COVID-19. Forest plot assessing the relative risk (RR) and 95% confidence interval (95% CI) of mortality in adults hospitalized with community-acquired and probable nosocomial COVID-19, according to the study definitions. The size of each box is proportional to the size of the individual hospital site (A-N), with the error bars representing the 95% CIs. The diamond represents the pooled average across studies, based on a random effects (RE) model. I^2^ : heterogeneity variance, calculated using restricted effects maximum likelihood (REML).

### 3.6 Meta-analysis of critical care admission

Critical care admission rates were reported in 8 studies reporting nosocomial outbreaks (11,13,34,39,42,44,45,47); with a crude rate of 27/252 (10.7%) in patients with nosocomial COVID-19 compared to 359/1396 (25.7%) in those hospitalised with community-acquired COVID-19. Meta-analysis is shown in Figure 4, with the pooled relative risk indicating this trend did not reach statistical significance (RR= 0.70, 95% CI: 0.48 to 1.03).

**Figure 4:**
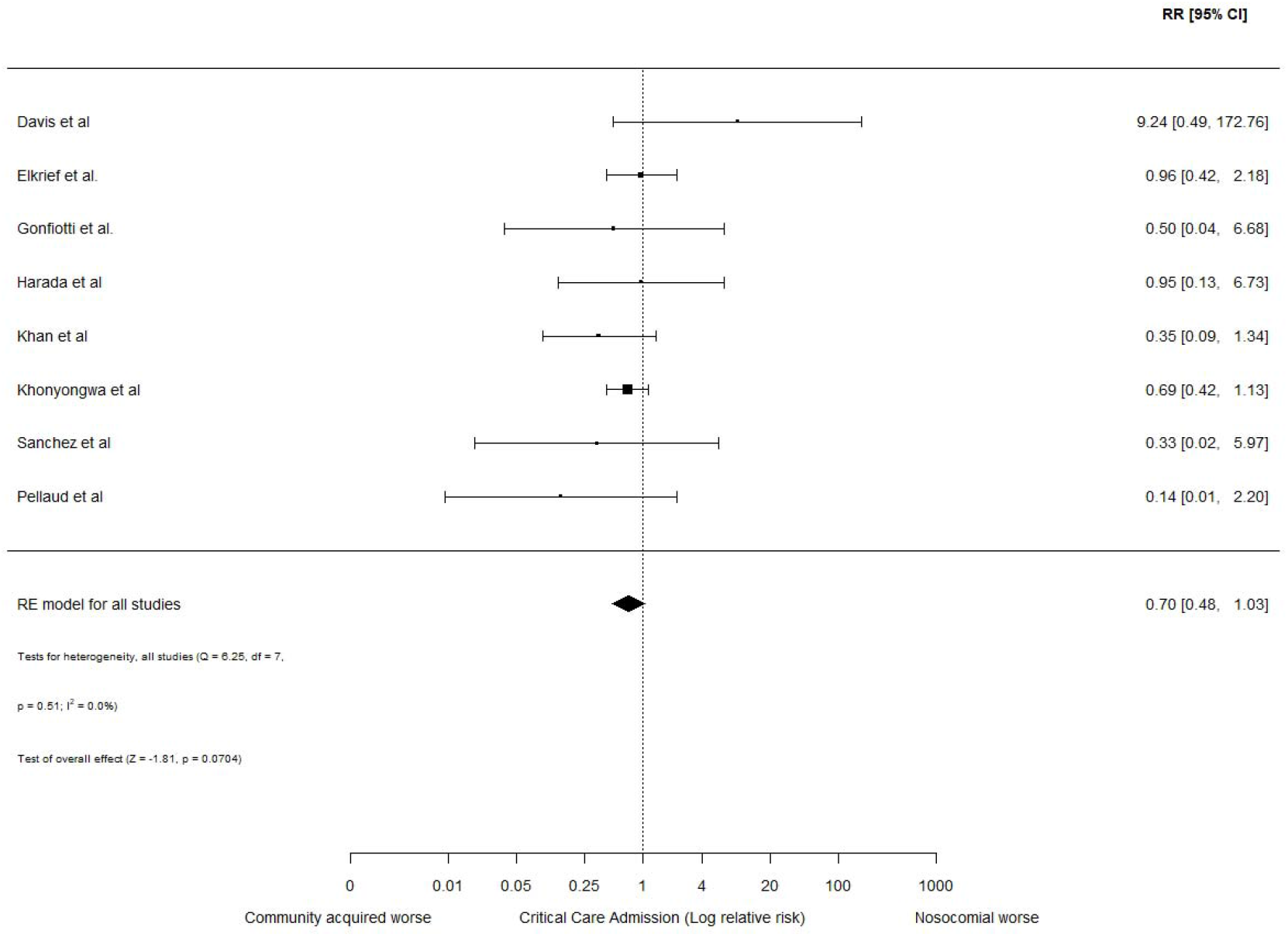
Relative risk of critical care admission in hospitalized adults with nosocomial and community-acquired COVID-19. Forest plot assessing the relative risk (RR) and 95% confidence interval (95% CI) of critical care admission in adults hospitalized with community-acquired and probable nosocomial COVID-19. The size of each box is proportional to the size of the individual hospital site (A-N), with the error bars representing the 95% CIs. The diamond represents the pooled average across studies, based on a random effects (RE) model. I^2^ : heterogeneity variance, calculated using restricted effects maximum likelihood (REML).

### 3.7 Sensitivity analysis

To challenge the robustness of our findings, we examined the effect of varying the level of certainty of nosocomial case diagnosis, study quality, and use of imputed mortality data across 5 sensitivity analyses, and assessed if individual studies conferred undue influence (Supplementary S3). These suggested that no individual study had undue influence on the results. Exclusion of studies across all sub-groups led to similar point estimates for the relative risk of mortality, but did not reach statistical significance in 4 of 5 assessments. Considering the immunosuppressed subgroup, the directionality and significance of our findings remained unchanged across 4 of 5 sensitivity analyses. Summary statistics for age were reported in 1291 nosocomial (mean 77.2 years), and 4542 community-acquired COVID-19 (mean 70.1 years) admissions. Gender was available in 1313 nosocomial (49.7% male) and 4375 (56.6% male) community-acquired COVID-19 cases. These differences precluded meta-regression analysis.

### 3.8 Reporting biases

We assessed for publication bias by examining the cumulative evidence distribution for our primary outcome using a funnel plot (Figure 5). Egger’s test did not suggest funnel plot asymmetry (p=0.51). Given the potentially sensitive implications of nosocomial infection (48), we hypothesised selective reporting of mortality might exist between nations. We therefore compared the frequency and origin of reports identified at the full text eligibility review stage meeting our study definition of a nosocomial outbreak (n= 67), with those including mortality as an outcome within this patient group independent of community outcomes. Overall 38 studies included mortality as an outcome (including 5 studies without observed nosocomial deaths), equating to a mortality reporting rate of 57%. Table 5 shows variation in the rate of mortality reporting by country. Reports from the UK accounted for 21/67 (31%) of nosocomial reports, and included mortality an outcome in 15/21 (71%). By contrast, reports from the United States contributed 7/67 (10%) of international reports describing nosocomial outbreaks, however none reported mortality as an outcome measure. This deviated significantly from the predicted international reporting rate (Fisher’s exact test, p = 0.0018). Together, this suggests publication bias may be present.

**Table 5:** Rates of mortality reporting in nosocomial COVID-19 outbreaks, by country of origin

| Country | Nosocomial outbreak reported |  | Nosocomial mortality reported as an outcome* |  |
| --- | --- | --- | --- | --- |
|  | Total studies, n | Total of studies (%) | Included studies, n | Fraction of countries' total reports |
| United Kingdom, UK | 21 | 31% | 15 | 71% |
| United States, US | 7 | 10% | 0 | 0% |
| China | 6 | 9% | 4 | 67% |
| Spain | 5 | 7% | 3 | 60% |
| France | 3 | 4% | 2 | 67% |
| Belgium | 3 | 4% | 0 | 0% |
| Italy | 3 | 4% | 2 | 67% |
| Switzerland | 3 | 4% | 1 | 33% |
| South Korea | 2 | 3% | 1 | 50% |
| Brazil | 2 | 3% | 2 | 100% |
| Japan | 2 | 3% | 2 | 100% |
| Vietnam | 2 | 3% | 0 | 0% |
| Germany | 2 | 3% | 1 | 50% |
| International | 1 | 1% | 1 | 100% |
| Poland | 1 | 1% | 1 | 100% |
| Denmark | 1 | 1% | 0 | 0% |
| India | 1 | 1% | 1 | 100% |
| Canada | 1 | 1% | 1 | 100% |
| Ireland | 1 | 1% | 1 | 100% |
| **Total** | **67** | **-** | **38** | **57%** |

**Figure 5:**
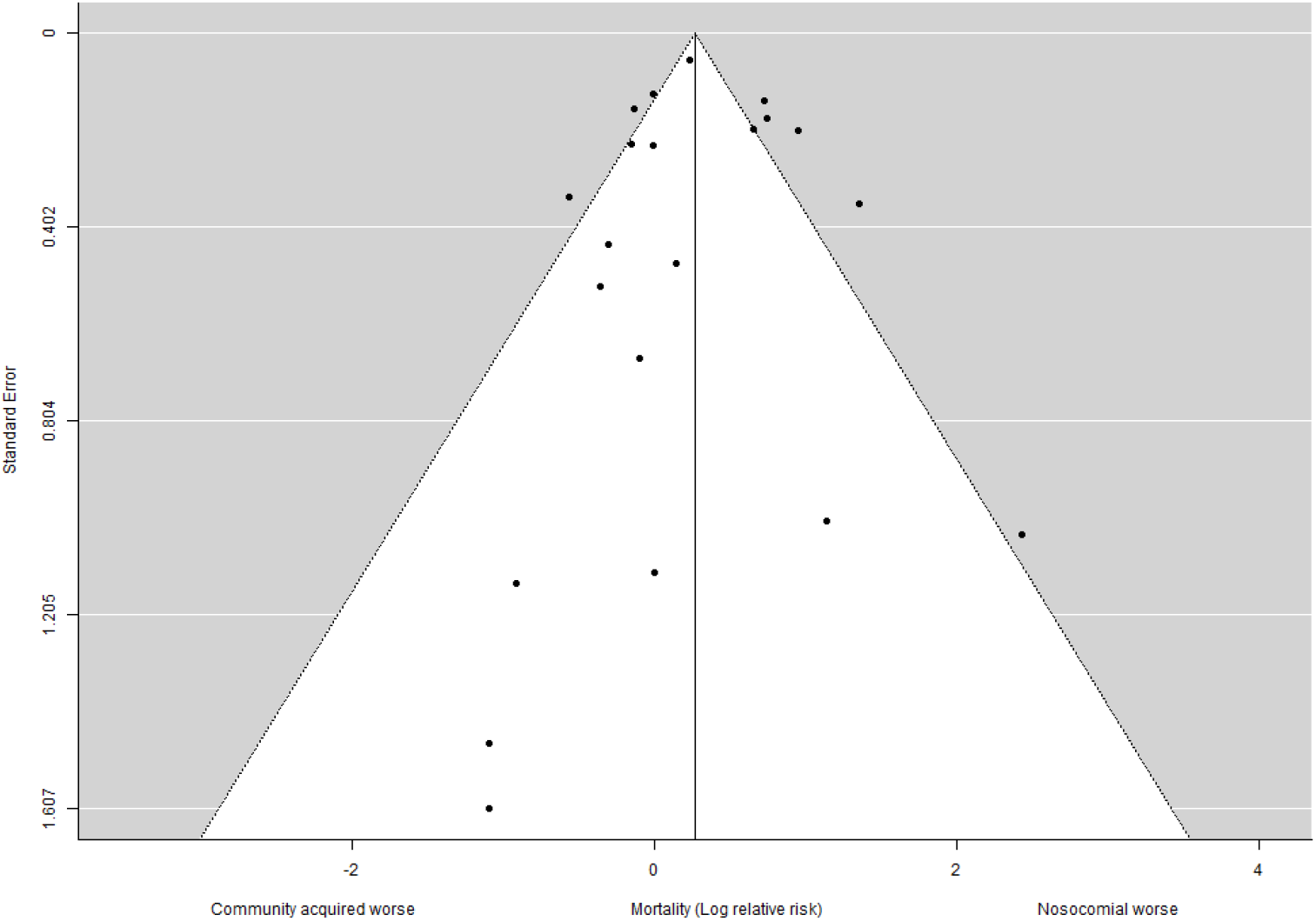
Funnel plot. Funnel plot with pseudo 95% confidence limits showing the distribution of relative risk of mortality across individual studies. Egger’s test, p=0.51.

### 3.9 Certainty of evidence

We assessed the quality of evidence supporting the statement: “In the general adult population, nosocomial COVID-19 is associated with a greater risk of inpatient mortality compared to individuals hospitalised with community-acquired COVID-19” as very low; and low/moderate in relation to “In an immunosuppressed adult population, nosocomial COVID-19 is associated with a greater risk of inpatient mortality compared to individuals hospitalised with community-acquired COVID-19”. Full GRADE assessment is shown in Table 6.

**Table 6:** Grading of Recommendations, Assessment, Development and Evaluations (GRADE) assessment.

| Statement | Number of studies and patients | Risk of bias | Indirectness | Inconsistency | Imprecision | Other considerations | Effect size | Overall quality of evidence |
| --- | --- | --- | --- | --- | --- | --- | --- | --- |
| “In the general adult population, nosocomial COVID-19 is associated with a greater risk of inpatient mortality compared to individuals hospitalised with community-acquired COVID-19” | 21 studies, 8246 patients.<br>Probable nosocomial: 1517<br>Probable community: 6729 | Serious - Very serious | Not serious | Very serious | Not serious | Publication bias suspected <sup>2</sup> | RR 1.31<br>95% CI: 1.01 to 1.70 | Low/ very low |
| “In an immunosuppressed adult population, nosocomial COVID-19 is associated with a greater risk of inpatient mortality compared to individuals hospitalised with community-acquired COVID-19” | 3 studies, 1069 patients.<br>Probable nosocomial: 152<br>Probable community: 917 | Serious* | Not serious | Not serious | Serious<sup>1</sup> | Publication bias suspected <sup>2</sup><br>Strong association <sup>3</sup> | RR 2.14<br>95% CI: 1.76 to 2.61 | Low/ Moderate |

Created using GRADEPro online tool, https://gradepror.org/. \* All studies scored moderate/high in formal assessment, however follow-up duration was limited; <sup>1</sup> Significant uncertainty associated with heterogeneity assessment: $I^2 = 0.00\%$ , 95% CI: 0.00 to 96.6%, downgrade by 1 level; <sup>2</sup> Mortality reporting bias suspected by country, downgrade by 1 level; <sup>3</sup> $RR > 2.0$ with consistent effect from $\geq 2$ studies, upgrade by 1 level.

## 4 Discussion

In this systematic review and meta-analysis addressing the burden of nosocomial COVID-19, we show the case fatality rate for nosocomial COVID-19 appears greater than community-acquired COVID-19, with a relative risk of 1.31 (95% CI: 1.01 to 1.70). Strikingly, we found that patients with malignancy (11,37) or transplant recipients (33) had approximately double the risk of dying after acquiring COVID-19 in hospital, compared to those hospitalised with community-acquired infection. This equates to a crude absolute mortality rate of 50.7% vs 23.8% respectively, with a consistent effect across studies and proved robust to sensitivity analyses assessing multiple assumptions around the certainty of nosocomial COVID-19 diagnosis and study quality.

The convergence of widely recognized risk factors for adverse outcomes in community-acquired COVID-19 in hospitalised patient groups, such as advanced age and frailty, are likely to contribute to the exaggerated mortality burden observed with nosocomial COVID-19. A range of potential mechanisms are likely to link individuals with cancer or recipients of transplants with mortality risk from nosocomial COVID-19, including both immunosuppression linked to the underlying condition and/or treatments and exposure due to health care requirements necessitating admission to the acute hospital environment. This is convergent with the heightened risk of mortality from COVID-19 independently reported for individuals with inherited and acquired forms of immunodeficiency (16), and the wider susceptibility of patients with haematological malignancy across a spectrum of healthcare-associated infections (49). Individual studies suggested a relationship between mortality rates and degree of immunosuppression, with the greatest mortality rate observed in patients with haematological malignancies who had recently received chemotherapy (37). This is consistent with results from patients enrolled within the UK Coronavirus Cancer Monitoring Project, which included 227 patients with haematological malignancies diagnosed with COVID-19. In this setting, recent chemotherapy approximately doubled the odds of dying during COVID-19-associated hospital admission (odds ratio: 2.09; 95% CI 1.09 to 4.08) after adjusting for age and gender; however, this study did not account for nosocomial infection.

Our study has several strengths. We systematically screened both the peer-reviewed and pre-print literature, leveraging the enhanced availability of full-texts by many publishers, to summarise the outcomes of 8246 adults hospitalised with COVID-19 during the first wave of the pandemic across 8 countries. This work establishes a relevant baseline for subsequent and future waves of the COVID-19 pandemic, and to our knowledge, represents the first meta-analysis of nosocomial COVID-19 mortality rates published to date. Zhou et al reported a rapid review and meta-analysis of nosocomial infections due to a range of viral pandemic threats, but included only 3 studies with SARS-CoV-2 and did not consider mortality as an outcome (7). In order to support the generalisability of our findings, we included studies with implicit and explicit definitions of nosocomial COVID-19. Accordingly, we catalogued a wide spectrum of case definitions, including combined epidemiological and genomic viral sequencing (32). We controlled for this variation in case definitions within our sensitivity analyses, for instance utilising outcomes meeting consensus international criteria for definite nosocomial infection wherever available. Although our funnel plot did not indicate publication bias amongst studies reporting mortality, our sequential literature review process suggests variation in the frequency of mortality reporting associated with studies describing nosocomial COVID-19 outbreaks. In particular, we identified no studies reporting mortality associated with nosocomial COVID-19 infection outbreaks originating from the United States, despite the high rate of COVID-19 cases and mortality in this country to date (50). Of the 7 studies we identified reporting nosocomial COVID-19 at the full text review stage, four dealt only with incidence (51–54), whilst three reported mortality but without reference to probable origin (55–57). Whilst we cannot exclude the risk of reporting bias, given the sensitive nature of this topic (48), this observation highlights successful infection control practices. Reporting on experience from a large US academic medical centre, Rhee et al found that despite a high burden of COVID-19, only two patients likely acquired COVID-19 during their admission (51). Generalising these practices may constitute a challenge across global health care settings acutely, for instance shortages of negative pressure isolation rooms were reported during the first wave in UK hospitals (44), but remain relevant as part of a longer-term “rebuild better” strategy.

Our study also has limitations, including its focus on hospitalised patients during the first wave of the pandemic. This is likely to introduce both selection and reporting bias, as during this period limited capacity meant RT-PCR testing was initially restricted to symptomatic individuals in the community (40,41). Estimates of age-stratified infection fatality rates in the adult UK general population during the first wave ranged from 0.03% (20-29 years) to 7.8% (over 80 years) (58), far lower than the inpatient comparator mortality rate used in our analysis. By contrast, individuals admitted during nosocomial outbreaks were more likely to be subject to screening, resulting in sampling of individuals across the true spectrum of disease severities (34,44), including earlier in their disease course. Our risk of bias assessment therefore focused on study inclusion and adequate follow-up as essential domains, to account for unequal disease progression at study entry between groups. Taken together, this considerably strengthens evidence for the statement: “nosocomial COVID-19 was associated with a greater risk of mortality, compared to individuals with community-acquired COVID-19 in the general adult population during the first wave of the COVID-19 pandemic”. Due to limitations in the summary data available, we were unable to perform planned meta-regression for factors such as age, gender, frailty, ethnicity or deprivation. Similarly, as studies typically reported all-cause mortality, we cannot exclude that deaths may have occurred that were not directly linked to COVID-19. However, our findings are supported by examination of COVID-19-linked mortality data within the United Kingdom (59,60), which indicate that COVID-19 multiplies the risk of death associated with various underlying diagnoses (59).

In conclusion, we systematically gathered data from the international literature and demonstrate an increased relative risk of mortality associated with nosocomial and community COVID-19. This maybe underestimated due to consideration of only hospitalised individuals. In particular, we strengthen observational evidence indicating individuals with malignancy or transplant recipients are at markedly elevated risk of death when infected by SARS-CoV-2 in hospital, compared to the community. Although focusing on the first wave, our findings are likely of ongoing significance given confirmation of an impaired SARS-CoV-2 vaccine response in multiple immunosuppressed patient groups (61–63), and the continued occurrence of new viral variants with enhanced transmissibility. Together, these findings inform policy makers by strongly advocating continued public health surveillance, stringent infection control measures (51), and access to individualised clinical interventions (64,65) to combat the threat of nosocomial COVID-19.

## Supporting information

Appendix- PRISMA checklist

## Data Availability

The datasets analysed for this study will be made available upon written request to the corresponding author.

## 6 Author Contributions

MJP conceived the project and drafted the protocol with TJCW and SS, with supervision from SB, SJ, IH, and DF. MJP, SS, TJCW, KO, CD, and DS screened abstracts and performed the full text review. MJP, TJCW and SS performed the data quality assessment. TJCW and MJP analysed the data. MJP prepared the first draft of the manuscript. All authors contributed to manuscript revision, read, and approved the submitted version.

## 8 Ethical approval

The data used in this work obtained relevant participant consent and ethical approval, therefore no additional approvals were required.

## 9 Patient and public involvement

No patients or participants were involved in setting the research question or the outcome measures, nor were they involved in developing plans for design or implementation of the study. No patients were asked to advise on interpretation or writing up of results. The published manuscript relating to this work will be sent to the UK Coronavirus Immunology Consortium (UKCIC) for appropriate dissemination to participants.

## 10 Supplementary Material

Supplementary Materials are available online.

## References

1. Burke JP. Infection Control — A Problem for Patient Safety. New England Journal of Medicine. 2003;348(7):651–6.

2. Allegranzi B, Nejad SB, Combescure C, Graafmans W, Attar H, Donaldson L, et al. Burden of endemic health-care-associated infection in developing countries: systematic review and meta-analysis. The Lancet. 2011;377(9761):228–41.

3. Senior K. Can we keep up with hospital-acquired infections? The Lancet Infectious Diseases. 2001;1:8.

4. Godoy P, Torner N, Soldevila N, Rius C, Jane M, Martínez A, et al. Hospital-acquired influenza infections detected by a surveillance system over six seasons, from 2010/2011 to 2015/2016. BMC Infectious Diseases. 2020 Jan 28;20(1):80.

5. NHS England and NHS Improvement, Chief Executives and Chief Operating Officers of all NHS Trusts and Foundation Trusts, CCG Accountable Officers. Minimising nosocomial infections in the NHS. 2020.

6. Bak A, Mugglestone MA, Ratnaraja NV, Wilson JA, Rivett L, Stoneham SM, et al. SARS-CoV-2 routes of transmission and recommendations for preventing acquisition: joint British Infection Association (BIA), Healthcare Infection Society (HIS), Infection Prevention Society (IPS) and Royal College of Pathologists (RCPath) guidance. Journal of Hospital Infection [Internet]. [cited 2021 Jun 24]; Available from: https://doi.org/10.1016/j.jhin.2021.04.027

7. Zhou Q, Gao Y, Wang X, Liu R, Du P, Wang X, et al. Nosocomial infections among patients with COVID-19, SARS and MERS: a rapid review and meta-analysis. Ann Transl Med. 2020;8(10):629–629.

8. PHE; LSHTM; The contribution of nosocomial infections to the first wave. Scientific Advisory Group for Emergencies (SAGE) [Internet]. 2021 Feb 12; Available from: https://www.gov.uk/government/publications/phe-and-lshtm-the-contribution-of-nosocomial-infections-to-the-first-wave-28-january-2021

9. Ponsford MJ, Jefferies R, Davies C, Farewell D, Humphreys IR, Jolles S, et al. The burden of nosocomial covid-19 in Wales: results from a multi-centre retrospective observational study of 2508 hospitalized adults. Thorax. 2021 Jun 18;manuscript in press.

10. Lee J. Y, Kim H. A, Huh K, Hyun M, Rhee J. Y, Jang S, et al. Risk Factors for Mortality and Respiratory Support in Elderly Patients Hospitalized with COVID-19 in Korea. J Korean Med Sci. 2020;35(23):e223.

11. Elkrief A, Desilets A, Papneja N, Cvetkovic L, Groleau C, Lakehal Y. A, et al. High mortality among hospital-acquired COVID-19 infection in patients with cancer: A multicentre observational cohort study. European Journal of Cancer. 2020;139:181–7.

12. Carter B, Collins JT, Barlow-Pay F, Rickard F, Bruce E, Verduri A, et al. Nosocomial COVID-19 infection: examining the risk of mortality. The COPE-Nosocomial Study (COVID in Older PEople). Journal of Hospital Infection. 2020;106(2):376–84.

13. Khan KS, Reed-Embleton H, Lewis J, Saldanha J, Mahmud S. Does nosocomial SARS-CoV-2 infection result in increased 30-day mortality? A multi-centre observational study to identify risk factors for worse outcomes in COVID-19 disease. J Hosp Infect. 2020 Sep 17;107:91–4.

14. Richterman A, Meyerowitz E. A, Cevik M. Hospital-Acquired SARS-CoV-2 Infection: Lessons for Public Health. JAMA - Journal of the American Medical Association. 2020;324(21):2155–6.

15. Williamson EJ, Walker AJ, Bhaskaran K, Bacon S, Bates C, Morton CE, et al. Factors associated with COVID-19-related death using OpenSAFELY. Nature. 2020 Aug 1;584(7821):430–6.

16. Shields AM, Burns SO, Savic S, Richter AG, Anantharachagan A, Arumugakani G, et al. COVID-19 in patients with primary and secondary immunodeficiency: The United Kingdom experience. Journal of Allergy and Clinical Immunology.

17. Khalid U, Ilham MA, Nagaraja P, Elker D, Asderakis A. SARS-CoV-2 in Kidney Transplant and Waitlisted Patients During the First Peak: The Welsh Experience. Transplant Proc. 2020/12/19 ed. 2021 May;53(4):1154–9.

18. Hewitt J, Carter B, Vilches-Moraga A, Quinn TJ, Braude P, Verduri A, et al. The effect of frailty on survival in patients with COVID-19 (COPE): a multicentre, European, observational cohort study. The Lancet Public Health. 2020;5(8):e444–51.

19. Richards SJG, D’Souza J, Pascoe R, Falloon M, Frizelle FA. Prevalence of frailty in a tertiary hospital: A point prevalence observational study. PLOS ONE. 2019;14(7):e0219083– e0219083.

20. Page MJ, Bossuyt PM, Boutron I, Hoffmann TC, Mulrow CD, Shamseer L, et al. The PRISMA 2020 statement: an updated guideline for reporting systematic reviews. BMJ. 2021 Mar 29;372:71.

21. Bandyopadhyay S, Baticulon RE, Kadhum M, Alser M, Ojuka DK, Badereddin Y, et al. Infection and mortality of healthcare workers worldwide from COVID-19: a systematic review. BMJ Global Health. 2020 Dec 1;5(12):e003097.

22. Abbas M, Robalo Nunes T, Martischang R, Zingg W, Iten A, Pittet D, et al. Nosocomial transmission and outbreaks of coronavirus disease 2019: the need to protect both patients and healthcare workers. Antimicrobial Resistance and Infection Control. 2021;10(1):7.

23. Ouzzani M, Hammady H, Fedorowicz Z, Elmagarmid A. Rayyan—a web and mobile app for systematic reviews. Systematic Reviews. 2016 Dec 5;5(1):210.

24. Moola S, Munn Z, Tufanaru C, Aromataris E, Sears K. Chapter 7: Systematic reviews of etiology and risk. In: Joanna Briggs Institute Reviewer’s Manual. https://reviewersmanual.joannabriggs.org/;

25. NICE. Appendix H: Appraisal checklists, evidence tables, GRADE and economic profiles. In: Developing NICE guidelines: the manual. (Process and methods [PMG20]).

26. Langan D, Higgins JPT, Jackson D, Bowden J, Veroniki AA, Kontopantelis E, et al. A comparison of heterogeneity variance estimators in simulated random-effects meta-analyses. Research synthesis methods. 2019 Mar;10(1):83–98.

27. Perera R, Heneghan C. Interpreting meta-analysis in systematic reviews. Evid Based Med. 2008 Jun 1;13(3):67.

28. Higgins JPT, Thomas J, Chandler J, Cumpston M, Li T, Page MJ WV. Cochrane Handbook for Systematic Reviews of Interventions. Cochrane. 2021;version 6.

29. Higgins JPT, Thompson SG, Deeks JJ, Altman DG. Measuring inconsistency in meta-analyses. BMJ. 2003 Sep 6;327(7414):557–60.

30. Guyatt GH, Oxman AD, Vist GE, Kunz R, Falck-Ytter Y, Alonso-Coello P, et al. GRADE: an emerging consensus on rating quality of evidence and strength of recommendations. BMJ. 2008;336(7650):924–6.

31. Vanhems P, Saadatian-Elahi M, Chuzeville M, Marion E, Favrelle L, Hilliquin D, et al. Rapid nosocomial spread of SARS-CoV-2 in a French geriatric unit. Infect Control Hosp Epidemiol. 2020;41(7):866–7.

32. Snell LB, Fisher CL, Taj U, Merrick B, Alcolea-Medina A, Charalampous T, et al. Combined epidemiological and genomic analysis of nosocomial SARS-CoV-2 transmission identifies community social distancing as the dominant intervention reducing outbreaks. medRxiv. 2020;2020.11.17.20232827.

33. Coll E, Fernandez-Ruiz M, Sanchez-Alvarez J. E, Martinez-Fernandez J. R, Crespo M, Gayoso J, et al. Covid-19 in transplant recipients: the spanish experience. American journal of transplantation_: official journal of the American Society of Transplantation and the American Society of Transplant Surgeons. 2020;

34. Davis P, Gibson R, Wright E, Bryan A, Ingram J, Lee RP, et al. Atypical presentations in the hospitalized older adult testing positive for SARS-CoV-2: a retrospective observational study in Glasgow, Scotland. Scott Med J. 2020 Oct 11;36933020962891.

35. Garatti A, Castelvecchio S, Daprati A, Molfetta R, Volpe M, De Vincentiis C, et al. Clinical Course of COVID-19 Infection in Patients Urgently Operated of Cardiac Surgical Procedures. Annals of surgery. 2020;272(4):e275–9.

36. Ajayi B, Trompeter A, Arnander M, Sedgwick P, Lui DF. 40 days and 40 nights: Clinical characteristics of major trauma and orthopaedic injury comparing the incubation and lockdown phases of COVID-19 infection. Bone & joint open. 2020;1(7):330–8.

37. Bhogal T, Khan U. T, Lee R, Stockdale A, Hesford C, Potti-Dhananjaya V, et al. Haematological malignancy and nosocomial transmission are associated with an increased risk of death from COVID-19: results of a multi-center UK cohort. Leukemia and Lymphoma. 2021;

38. Brill SE, Jarvis HC, Ozcan E, Burns TLP, Warraich RA, Amani LJ, et al. COVID-19: a retrospective cohort study with focus on the over-80s and hospital-onset disease. BMC Med. 2020 Jun 25;18(1):194.

39. Gonfiotti A, Gatteschi L, Salvicchi A, Bongiolatti S, Lavorini F, Voltolini L. Clinical courses and outcomes of five patients with primary lung cancer surgically treated while affected by Severe acute respiratory syndrome coronavirus 2. European journal of cardio-thoracic surgery_: official journal of the European Association for Cardio-thoracic Surgery. 2020;58(3):598–604.

40. Jewkes SV, Zhang Y, Nicholl DJ. Nosocomial spread of COVID-19: Lessons learned from an audit on a stroke/neurology ward in a UK district general hospital. Clinical Medicine, Journal of the Royal College of Physicians of London. 2020;20(5):E173–7.

41. Wake RM, Morgan M, Choi J, Winn S. Reducing nosocomial transmission of COVID-19: implementation of a COVID-19 triage system. Clin Med (Lond). 2020 Sep;20(5):e141–5.

42. Harada S, Uno S, Ando T, Iida M, Takano Y, Ishibashi Y, et al. Control of a Nosocomial Outbreak of COVID-19 in a University Hospital. Open Forum Infect Dis. 2020 Dec;7(12):ofaa512.

43. Cao J, Tu W. -J, Cheng W, Yu L, Liu Y. -K, Hu X, et al. Clinical features and short-term outcomes of 102 patients with coronavirus disease 2019 in Wuhan, China. Clinical Infectious Diseases. 2020;71(15):748–55.

44. Khonyongwa K, Taori SK, Soares A, Desai N, Sudhanva M, Bernal W, et al. Incidence and outcomes of healthcare-associated COVID-19 infections: significance of delayed diagnosis and correlation with staff absence. J Hosp Infect. 2020 Dec;106(4):663–72.

45. Sanchez M. D, Sanchez M, De La Morena J. M, Ogaya-Pinies G, Mateo E, Moscatiello P, et al. Nosocomial SARS-CoV-2 infection in urology departments: Results of a prospective multicentric study. International Journal of Urology. 2021;28(1):62–7.

46. Lakhani K, Minguell J, Guerra-Farfan E, Lara Y, Jambrina U, Pijoan J, et al. Nosocomial infection with SARS-CoV-2 and main outcomes after surgery within an orthopaedic surgery department in a tertiary trauma centre in Spain. International Orthopaedics. 2020;44(12):2505–13.

47. Pellaud C, Grandmaison G, Thien H. P.P.H, Baumberger M, Carrel G, Ksouri H, et al. Characteristics, comorbidities, 30-day outcome and in-hospital mortality of patients hospitalized with COVID-19 in a Swiss area - A retrospective cohort study. Swiss Medical Weekly. 2020;150(29):w20314.

48. Barranco R, Du Tremoul Lvb, Ventura FA. Hospital-acquired sars-cov-2 infections in patients: Inevitable conditions or medical malpractice? International Journal of Environmental Research and Public Health. 2021;18(2):1–9.

49. Liu H, Zhao J, Xing Y, Li M, Du M, Suo J, et al. Nosocomial Infection in Adult Admissions with Hematological Malignancies Originating from Different Lineages: A Prospective Observational Study. PLOS ONE. 2014 Nov 21;9(11):e113506.

50. Johns Hopkins Center for Systems Science and Engineering. The Johns Hopkins Coronavirus Resource Center (CRC).

51. Rhee C, Baker M, Vaidya V, Tucker R, Resnick A, Morris CA, et al. Incidence of Nosocomial COVID-19 in Patients Hospitalized at a Large US Academic Medical Center. JAMA Network Open. 2020;3(9):e2020498.

52. Long D. R, O’Reilly-Shah V, Rustagi A. S, Bryson-Cahn C, Jerome K. R, Weiss N. S, et al. Incidence of health care-associated COVID-19 during universal testing of medical and surgical admissions in a large US health system. Open Forum Infectious Diseases. 2020;7(10).

53. Thompson JWJ, Mikolajewski AJ, Kissinger P, McCrossen P, Smither A, Chamarthi GD, et al. An Epidemiologic Study of COVID-19 Patients in a State Psychiatric Hospital: High Penetrance With Early CDC Guidelines. Psychiatric services (Washington, DC). 2020;71(12):1285–7.

54. Nalleballe K, Siddamreddy S, Kovvuru S, Veerapaneni P, Roy B, Onteddu S. R. Risk of COVID-19 from hospital admission during the pandemic. Infect Control Hosp Epidemiol. 2020;

55. Mani V. R, Kalabin A, Valdivieso S. C, Murray-Ramcharan M, Donaldson B. New York inner city hospital COVID-19 experience and current data: Retrospective analysis at the epicenter of the American coronavirus outbreak. Journal of Medical Internet Research. 2020;22(9):e20548.

56. Dowlati E, Zhou T, Sarpong K, Pivazyan G, Briscoe J, Fayed I, et al. Case Volumes and Perioperative Coronavirus Disease 2019 Incidence in Neurosurgical Patients During a Pandemic: Experiences at Two Tertiary Care Centers in Washington, DC. World Neurosurgery. 2020 Nov;143:e550–60.

57. Hu P, Jansen J. O, Uhlich R, Black J, Pierce V, Hwang J, et al. Early comprehensive testing for COVID-19 is essential to protect trauma centers. Journal of Trauma and Acute Care Surgery. 2020;89(4):698–702.

58. Verity R, Okell LC, Dorigatti I, Winskill P, Whittaker C, Imai N, et al. Estimates of the severity of coronavirus disease 2019: a model-based analysis. The Lancet Infectious Diseases. 2020;20(6):669–77.

59. Bhaskaran K, Bacon S, Evans SJ, Bates CJ, Rentsch CT, MacKenna B, et al. Factors associated with deaths due to COVID-19 versus other causes: population-based cohort analysis of UK primary care data and linked national death registrations within the OpenSAFELY platform. The Lancet Regional Health – Europe [Internet]. 2021 Jul 1 [cited 2021 Jun 24];6. Available from: https://doi.org/10.1016/j.lanepe.2021.100109

60. Salmon R. L, Monaghan S. P. AO - Salmon R.L, ORCID: http://orcid.org/---. Who is dying from COVID-19 in the United Kingdom? A review of cremation authorisations from a single South Wales’ crematorium. Epidemiology and Infection. 2021;

61. Boyarsky BJ, Werbel WA, Avery RK, Tobian AAR, Massie AB, Segev DL, et al. Antibody Response to 2-Dose SARS-CoV-2 mRNA Vaccine Series in Solid Organ Transplant Recipients. JAMA. 2021 Jun 1;325(21):2204–6.

62. Parry HM, McIlroy G, Bruton R, Ali M, Stephens C, Damery S, et al. Antibody Responses After First and Second COVID-19 Vaccination in Patients With Chronic Lymphocytic Leukaemia. 2021; Available from: http://europepmc.org/abstract/PPR/PPR348446

63. Wadei HM, Gonwa TA, Leoni JC, Shah SZ, Aslam N, Speicher LL. COVID_19 infection in solid organ transplant recipients after SARS_CoV_2 vaccination. American journal of transplantation. 2021;

64. Kamar N, Abravanel F, Marion O, Couat C, Izopet J, Del Bello A. Three Doses of an mRNA Covid-19 Vaccine in Solid-Organ Transplant Recipients. N Engl J Med [Internet]. 2021 Jun 23 [cited 2021 Jun 25]; Available from: https://doi.org/0.1056/NEJMc2108861

65. Horby, W P, Mafham M, Peto L, Campbell M, Pessoa-Amorim G, et al. Casirivimab and imdevimab in patients admitted to hospital with COVID-19 (RECOVERY): a randomised, controlled, open-label, platform trial. medRxiv [Internet]. 2021; Available from: https://www.medrxiv.org/content/early/2021/06/16/2021.06.15.21258542

