## Supplementary Materials for "A systematic review and meta-analysis of inpatient mortality associated with nosocomial and community COVID-19 exposes the vulnerability of immunosuppressed adults"

### Supplementary online material

#### Supplementary S1: Extended Methods

Individual study variance and heterogeneity variance were combined to calculate individual study weight: $w_i = 1/(\tau^2 + \sigma^2)$. Studies which reported no observed deaths in both community acquired or nosocomial COVID-19 groups were excluded from the analysis. When only one group reported no deaths, a figure of 0.5 deaths was used for the purpose of analysis. Assumption of normality for meta-analysis models was assessed using Q-Q plots. To establish whether an individual study had undue influence on the meta-analysis model, the 'influence' function in the R metafor package was used. Studies were judged influential if one of the following was true:

- The absolute DFFITS value is larger than $3\sqrt{p/(k-p)}$, where p is the number of model coefficients and k the number of studies.
- The lower tail area of a chi-square distribution with p degrees of freedom cut off by the Cook's distance is larger than 50%.
- The hat value is larger than 3(p/k).
- Any DFBETAS value is larger than 1.

We pre-specified the following sensitivity analysis:

- 1: Studies providing an explicit definition of nosocomial acquisition
- 2: Studies providing outcomes associated with a standardised >14 day definition for 'definite' nosocomial covid-19 (excluding probable cases).
- 3A: Excluding studies with a higher risk of bias, defined as studies with a score of 4 or less.
- 3B: Fulfilling all 5 core study quality domains (as indicated by * within tables 2-4).
- 4: Excluding studies with imputed data (i.e. 0.5 used in place of zero-count cells)

#### Supplementary S2: Timing of non-UK studies included within primary meta-analyses relative to national COVID-19 rates

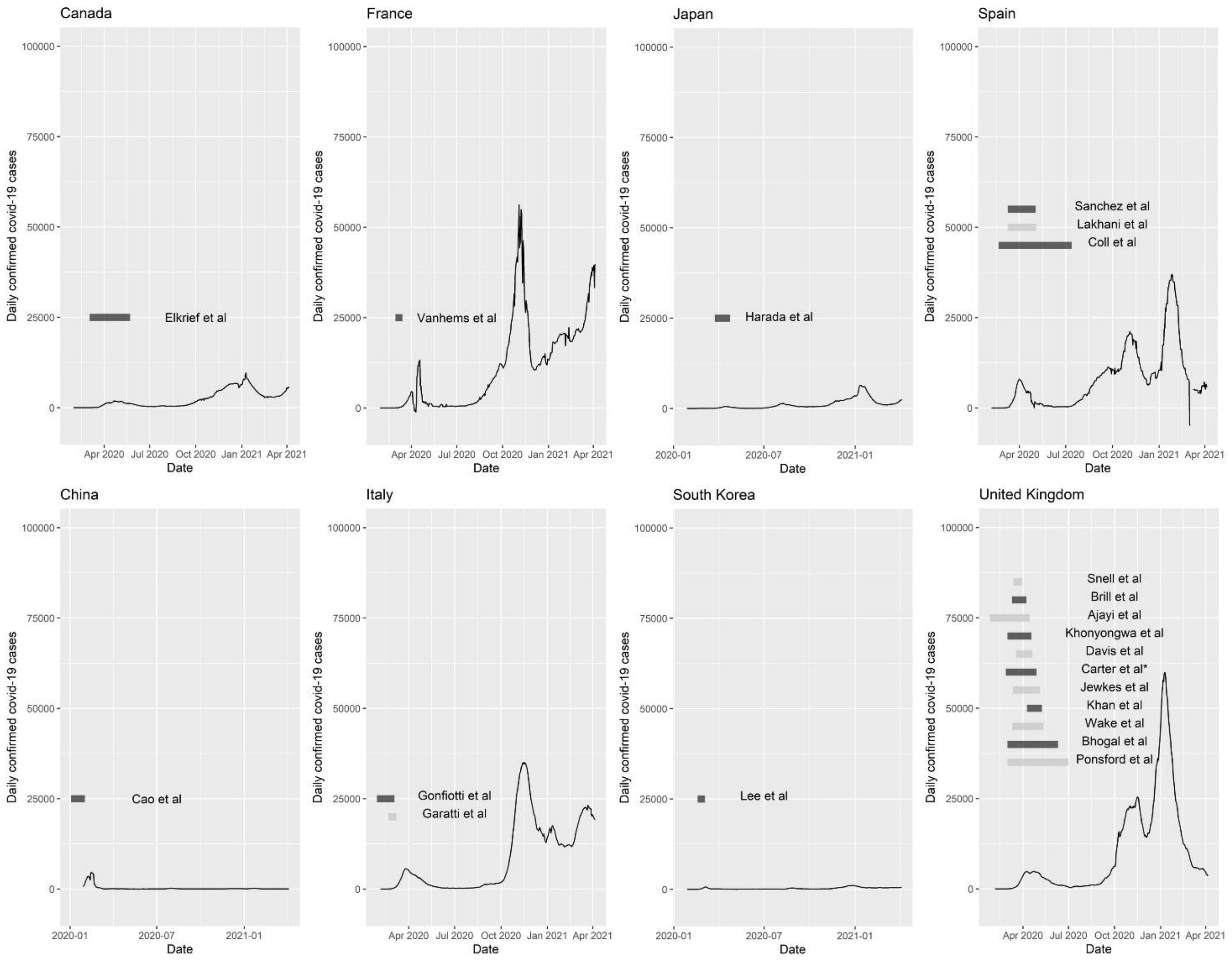

#### Supplementary S3: Sensitivity Analyses

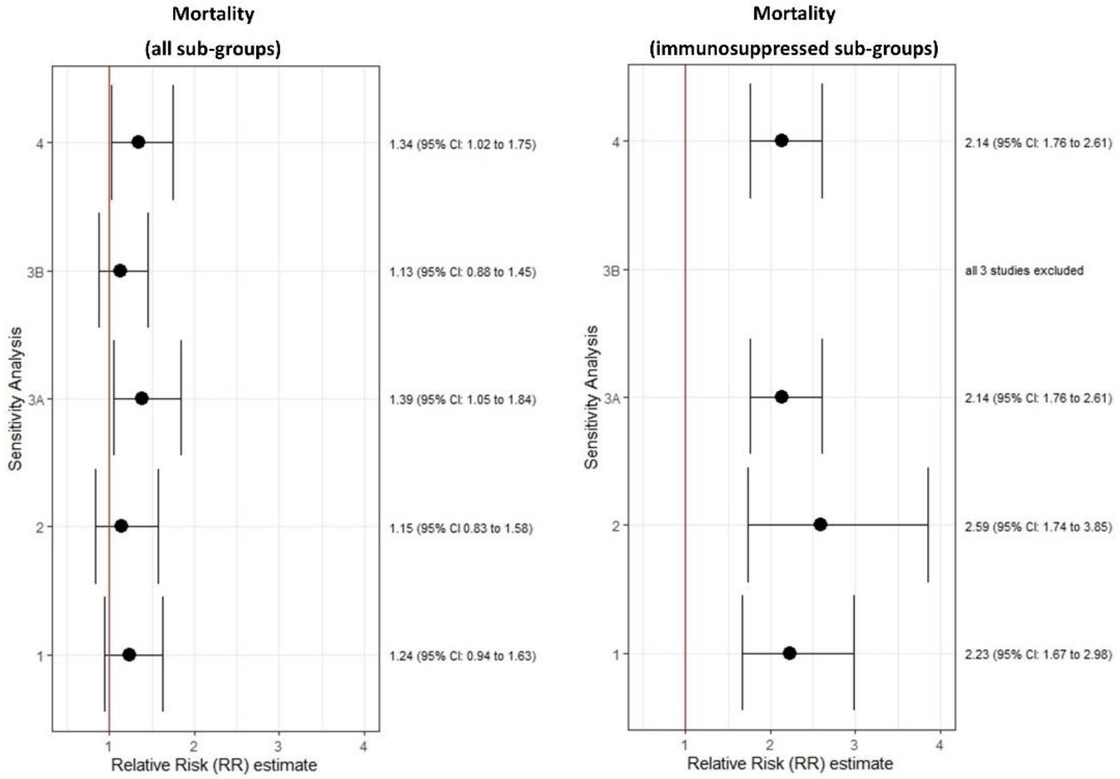

Forest plots showing the relative risk (RR, log estimate) and 95% confidence interval (95% CI) estimates for the relative risk of mortality in adults hospitalised with community-acquired and probable nosocomial COVID-19 applying pre-defined sensitivity analyses across subgroups or to immunosuppressed sub-group only. **1**: Studies providing an explicit definition of nosocomial acquisition; **2**: Studies providing outcomes associated with a standardized >14 day definition for 'definite' nosocomial covid-19; **3A**: Excluding studies with a higher risk of bias (indicated by total quality score <5); **3B**: Fulfilling all 5 core study quality domains (indicated by * within tables 2-4, see main article); **4**: Excluding studies with imputed data (i.e. 0.5 used in place of zero-count cells).

There was no significant difference in the analysis when studies without an explicit definition of nosocomial acquisition were excluded (n=19, p=0.78), however, removing these studies changed the overall relative risk for the difference between mortality from nosocomial and community acquired COVID-19 such that it no longer met the pre-defined 5% significance level (RR = 1.24, 95% CI 0.94 to 1.63, p= 0.13). There was no significant difference in the analysis when only studies providing outcomes based on standardised >14 day definition for 'definite' nosocomial covid-19 were included (n = 8, p= 0.54), however the difference between mortality from nosocomial and community acquired COVID-19 no longer met the pre-defined 5% significance level (RR = 1.15, 95% CI 0.83 to 1.58, p= 0.40). There was no significant change in mortality outcome when studies with a with "raw" risk of bias score of 5 or less were excluded (n=4, RR = 1.39 vs 1.31, p=0.76). There was no significant difference in the analysis when studies with high risk of bias were excluded (defined as studies scoring across all key 5 domains, indicated by * in Tables 2-4), n=12, p=0.42, however the difference between mortality from nosocomial and community acquired COVID-19 no longer met the pre-defined 5% significance level (RR = 1.13, 95% CI 0.88 to 1.45, p = 0.34). Excluding studies where data was imputed (n=3) had no significant effect on the results (RR 1.34 vs 1.31 p=0.91).
